# CHIP is associated with cardiovascular disease in the UK Biobank

**DOI:** 10.1101/2023.11.30.23299001

**Authors:** Caitlyn Vlasschaert, Giulio Genovese, Yash Pershad, Siddhartha Jaiswal, Pradeep Natarajan, Alexander G. Bick

**Affiliations:** Department of Medicine, Queen’s University, Kingston, Ontario, Canada; Program in Medical and Population Genetics, Broad Institute of MIT and Harvard, Cambridge, MA, USA; Stanley Center for Psychiatric Research, Broad Institute of MIT and Harvard, Cambridge, MA, USA; Department of Genetics, Harvard Medical School, Boston, MA, USA; Division of Genetic Medicine, Department of Medicine, Vanderbilt University Medical Center, Nashville, TN, USA; Department of Pathology, Stanford University School of Medicine, Stanford, CA, USA; Cardiovascular Research Center and Center for Genomic Medicine, Massachusetts General Hospital, Boston, MA, USA; Department of Medicine, Harvard Medical School, Boston, MA, USA

## Abstract

Clonal hematopoiesis (CH) is a form of age-related somatic mosaicism. CH encompasses both clonal events with recognizable leukemic driver mutations, such as CH of indeterminate potential (CHIP) and mosaic chromosomal alterations (mCAs), as well as clonal states without clear driver mutations. Stacey *et al*. identify cases of CH from whole genome sequencing (WGS) data in a subset of 130,709 UK Biobank (UKB) participants and in 45,510 individuals from an Icelandic cohort. They report that CH is not associated with cardiovascular disease (CVD) and posit that the multiple prior CH studies did not fully account for smoking-related confounding. We find that the conclusion reached by Stacey *et al*. is specific to their particular CH definition which groups well-established and clinically meaningful CH subtypes together despite evidence from multiple previous reports that distinct forms of CH have distinct phenotypic consequences. We show that (1) the CHIP/ CVD association in the UK Biobank is not confounded by smoking and (2) CH subtypes which Stacey *et al*. have lumped together have heterogenous associations with CVD. We suggest that Stacey et al.’s failure to identify an association between CHIP and CVD is perhaps related to methodologic differences compared to prior reports.

---

Clonal hematopoiesis (CH) is a form of age-related somatic mosaicism that confers risk of blood cancer and multiple other diseases. Since the phenomenon was first recognized decades ago there have been distinct methods for identifying clonality. For example, the identification of a cancer-associated driver mutation in an appreciable number of blood or marrow cells is *de facto* evidence for a clonal expansion event^1,2^. Other approaches can identify clonality even in the absence of a known driver, such as imbalanced X-chromosome inactivation in females^3^ or the detection of passenger mutations from sequencing of bulk DNA.^4^ Recently, Stacey *et al*. used an evolution of their previous method^5^ for identifying clonality in identify cases of CH from whole genome sequencing (WGS) data – itself an evolution of the method presented by Genovese *et al*.^4^ – in a subset of 130,709 UK Biobank (UKB) participants and in 45,510 individuals from their Icelandic cohort.^6^ Notably, they report that CH was not associated with cardiovascular disease (CVD) risk after adjusting for smoking, neither when CH was identified using their “Barcode-CH” method nor when CH was limited to candidate preleukemic driver (CPLD) mutations. They posit that previous positive reports^7–9^ were due to smoking-related confounding even though such reports adjusted for smoking and, in many studies, the effect size for CH was greater than that for smoking. We find that the conclusion reached by Stacey *et al*. is specific to their particular choice of CH definition which lumps well established and clinically meaningful CH categories together despite evidence from multiple previous reports that these distinct forms of CH have distinct phenotypic consequences.^1,10,11^

CH is a heterogeneous term that encompasses both clonal events with recognizable leukemic driver mutations as well as clonal states without clear driver mutations. Driver mutations can include point mutations or short insertions/deletions in myeloid leukemia driver genes such as *DNMT3A, TET2, ASXL1* and *JAK2* referred to as clonal hematopoiesis of indeterminate potential (CHIP) and mosaic chromosomal abnormalities (mCAs), which include both autosomal copy number changes and acquired loss of sex chromosomes^10,12^. Stacey *et al*. develop a CPLD definition that encompasses both the classic CHIP genes as well as other drivers. Distinct driver genes have different phenotypic associations, with driver genes such as *TET2* and *JAK2* particularly associated with CVD while *DNMT3A*, the most common CHIP driver gene, has more modest effects.^7,9^ Proposed mechanisms center around dysregulated inflammatory signaling by affected myeloid cells leading to endothelial injury, plaque instability, cellular infiltration, and myocardial fibrosis.^7,8^ In contrast, most mCAs have not been associated with CVD nor with inflammatory markers, with rare exceptions.^11^ Given that non-CPLD CH outnumbers CPLD CH (and classical CHIP) in Stacey *et al*., CH as a heterogeneous grouping is not expected to be associated with CVD.

When we examine CHIP and incident cardiovascular disease using our WES-derived CHIP call set in the 451,180-person UKB dataset^13^ and our published CVD phenotype^9,14^, we do observe a significant association in a model that adjusts for the same covariates used in Stacey *et al*. Table 2: linear and quadratic age, sex, and smoking status (**Figure 1A**). The CHIP/CVD association is stronger in non-*DNMT3A* genes. Importantly, CHIP and non-*DNMT3A* CHIP are associated with incident CVD in never-smokers with similar point estimates as compared with present and past smokers, suggesting that the CHIP signal is not the result of residual smoking-related confounding. Moreover, previously reports indicate that smoking is an uncommon comorbidity of CHIP, but is causally implicated in the development of mCAs.^15^ As previously reported by multiple groups^16–18^, mCA CH subtypes are not associated with incident CVD (**Figure 1B**). In addition, mutations in *TET2* have been most consistently associated with numerous cardiovascular outcomes, such as increased risk of coronary heart disease, heart failure, and stroke.^7,19–22^ We note that Stacey *et al*. found no association between mutations in *TET2* and smoking, consistent with prior data^20,23^, refuting their argument that the association of CH and CVD outcomes is due to residual confounding.

**Figure 1.**
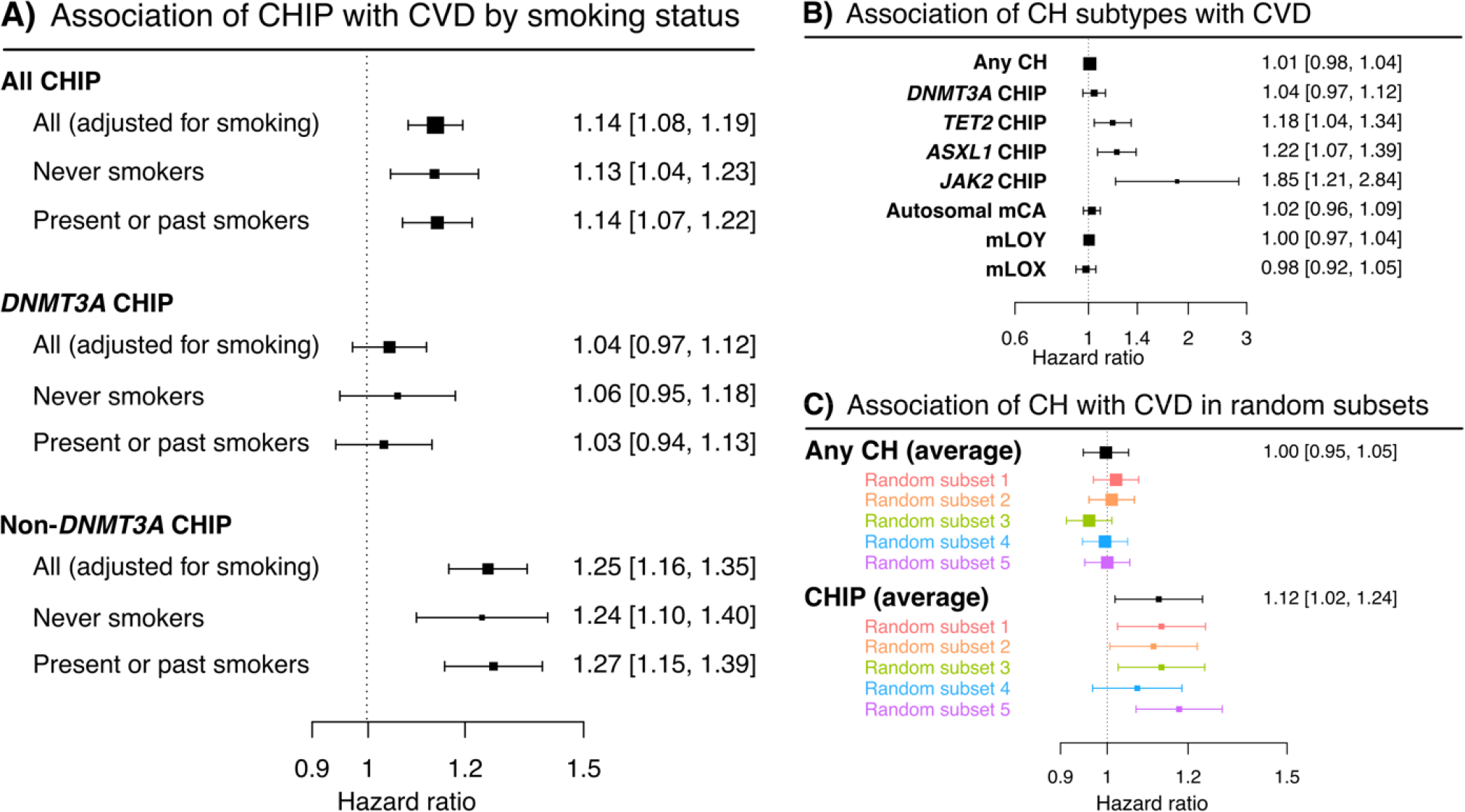
CHIP is associate with cardiovascular disease in the UK Biobank. **A)** CHIP and non-*DNMT3A* CHIP are equally associated with incident CVD in smokers and never-smokers in the full UK Biobank dataset. **B)** CH as umbrella classification is not associated with CVD as it includes autosomal and sex chromosome mCAs. **C)** CHIP is associated with CVD in random subsets of the UK Biobank, suggesting the lack of association reported by Stacey *et al*. is not due to statistical power but possibly related to differences in CHIP ascertainment. Consistent with Stacey *et al*., all analyses are adjusted for linear and quadratic age, sex, and binary smoking status (present or past smoking = 1, never smoking = 0).

It should be noted that CH as well as the CHIP and mLOY subtypes are associated with CVD when only large clones are considered in our analyses (**Supplemental Figure 1**), suggesting that clone size is an important consideration when evaluating CH associations in addition to CH subtype. Additionally, specific subtypes of autosomal mCAs have been associated with CVD, such as copy-neutral loss of heterozygosity of the p arm of chromosome 9 (9p-CNLOH).^24^

When we aggregate distinct classes of CH together, we obtain the same point estimate for “Any CH” as Stacey *et al*. do for their Barcode-CH analyses (HR 1.01, 95% CI: 0.98–1.04). While Stacey *et al*. report null findings when applying our previously described method to identify CHIP in a 118,673-person subset of the UKB with WGS data^13^ (HR 1.01, 95% CI: 0.88-1.15), we find positive associations for CHIP with CVD both in the full WES cohort using our CHIP calls and in randomly selected subsets of 118,673 individuals from the WES dataset (**Figure 1C**), suggesting their discordant findings are a result of their method to identify CHIP rather than statistical power. Notably, they selected a minimum allele depth (minAD) threshold of 3. We have previously identified the minAD as a critical determinant of CHIP dataset specificity and recommended that this parameter be systematically evaluated to determine the optimal threshold.^9,13^ We also note that the WES sequencing depth was greater than WGS, which may also contribute to differences in sensitivity to detect somatic mutations. It is also possible that the 118,673-person subset of the UKB WGS is not representative of the entire cohort. In summary, using <30% of the UKB participants with available genetic data, the authors obtain null results, whereas when we evaluate this association in both the full dataset and in random data subsets of 118,673 individuals using *bona fide* CHIP calls, we do observe an association between CHIP and CVD including in never smokers. Similar results identifying an association between CHIP and CVD in the full UK Biobank data set have been previously reported in an independent analysis by Kessler *et al*.^14^

Lastly the authors offer another possible explanation for their null findings: they note that they performed “stringent exclusion of people with a pre-existing hematologic abnormality” by excluding individuals who developed a hematologic malignancy within 6 months of enrollment. However, we and others have used this same exclusion criterion in our current and prior work, including when evaluating the role of CHIP in CVD.^9,13^

In conclusion, agglomeration of heterogenous kinds of CH together decreases statistical power for scientific discovery and impairs interpretability for clinical risk stratification. In this study we showed that the association between CHIP – a distinct category of CH – and CVD is robust, driven by specific driver genes and not confounded by smoking in the UK Biobank. We recommend that future CH-disease association studies focus on specific CH driver events to accelerate biological understanding and advance clinical utility.

## Data Availability

UK Biobank data is available to all registered researchers.

## Author contributions statement

C.V., S.J., P.N., and A.G.B. conceived the study. C.V., G.G., and Y.P. performed analyses. C.V. and A.G.B. drafted the manuscript, which was revised with critical input from all authors.

**Supplemental Figure 1.**
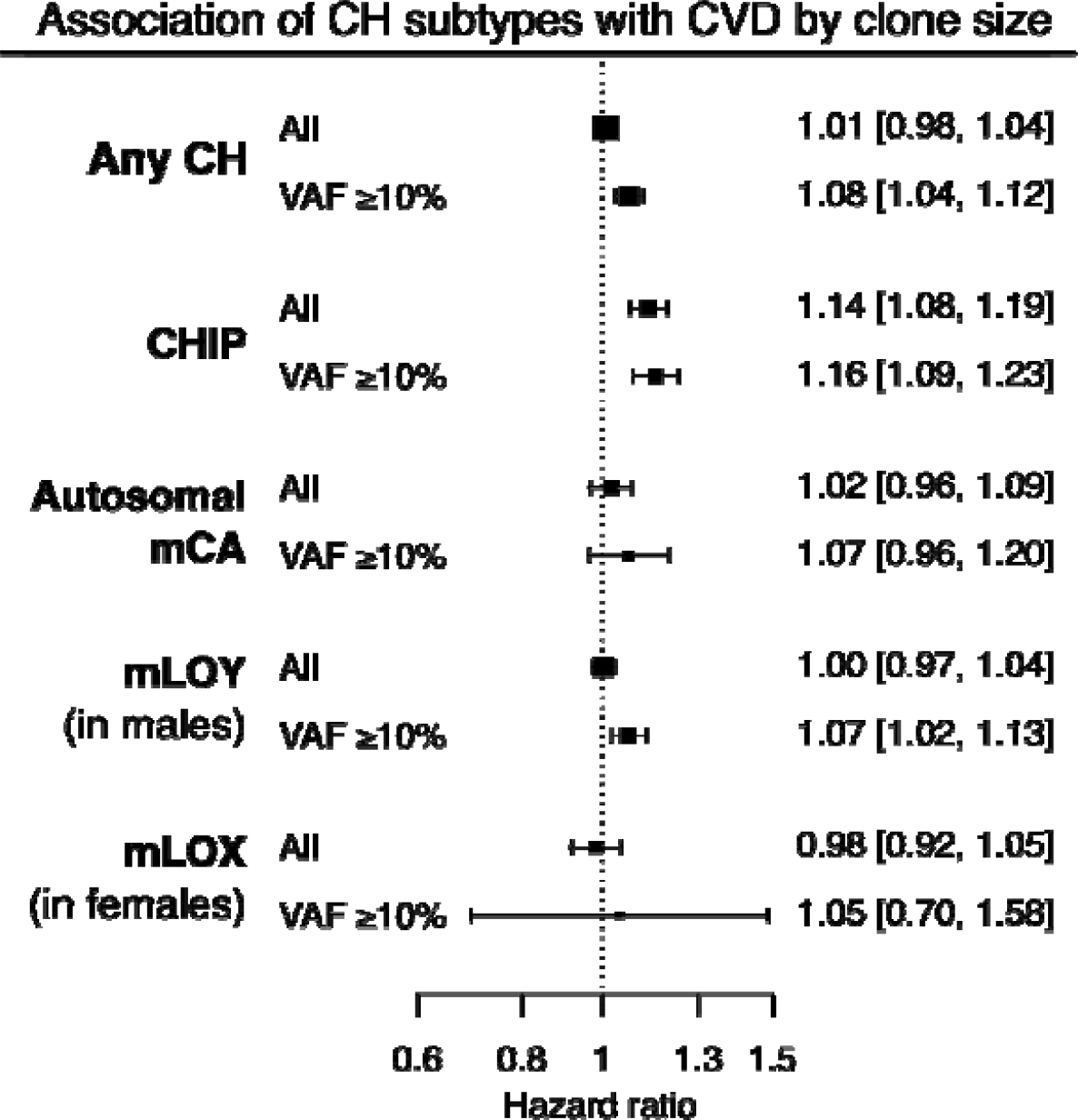
Large CH clones are associated with incident CVD. Note that a CHIP variant allele fraction (VAF) of 10% for diploid cells equates to a cell fraction (CF) of 20%.

